# FcγRIIIb-deficient neutrophils have defects in ROS production, phagocytosis and actin polymerization following stimulation through FcγRs

**DOI:** 10.1101/2025.05.08.25327271

**Authors:** José Antonio Cruz-Cárdenas, Jorge Andrés Cázares-Preciado, Alejandra López-Arredondo, Ana Beatriz Sánchez-Argáez, Michael Schnoor, Marion E.G. Brunck

**Affiliations:** Escuela de Ingeniería y Ciencias, Tecnológico de Monterrey, Monterrey, Nuevo León, México; Department of Molecular Biomedicine, CINVESTAV-IPN, Av IPN 2508, San Pedro Zacatenco, 07360 Mexico City, Mexico; The Institute for Obesity Research, Tecnológico de Monterrey, Monterrey, Nuevo León, México

**Keywords:** Neutrophils, FcγRIIIb, CD16b, Phagocytosis

## Abstract

Neutrophils are crucial to innate immune responses to microbes. The crosslinking of opsonized pathogens by Fc gamma receptors (FcγRs) on neutrophil surfaces mediates multiple antimicrobial functions, including phagocytosis and the production of reactive oxygen species (ROS). FcγRIIIb (CD16b) is the most abundant receptor on human neutrophils. It is a GPI-anchored receptor that lacks an intracellular domain. The exact mechanisms by which FcγRIIIb transduces signals remain unclear. A rare FcγRIIIb-deficient phenotype has been reported in apparently healthy subjects, which is intriguing given the abundance of this receptor on neutrophil surfaces and its crucial role in neutrophil activation by immune complexes. Here, we identified 2 healthy brothers lacking FcγRIIIb on neutrophils and characterized their neutrophil activation through FcγR crosslinking by immune complexes. Sequencing of the FCGR3B gene revealed mutations in exon 2 resulting in translation loss. In the absence of stimulation, FcγRIIIb**^null^** neutrophils showed unaltered levels of FcγRIIa, TLR-2, TLR-4 and TLR-6, but significantly higher FcγRIIIa and FcγRIa. Upon challenge with E. coli immune complexes, increased surface expression of FcγRIa, TLR-4, and αM integrin (CD11b) was observed exclusively in FcγRIIIb**^null^** neutrophils. Antibacterial functions stimulated by immune complexes were significantly lower in FcγRIIIb**^null^** neutrophils, including phagocytic capacity and ROS production compared to FcγRIIIb-expressing neutrophils. Overall, the absence of FcγRIIIb on human neutrophils correlated with impaired antimicrobial functions following stimulation through FcγRs. This study provides new insights into the functional relevance of FcγRIIIb and emphasizes the importance of this receptor in neutrophil responses to bacteria.

## Introduction

Fc gamma receptors (FcγR) on the surface of neutrophils crosslink IgG-opsonized microbes, prompting effector functions such as ROS production and phagocytosis that often culminate in pathogen clearance^1–3^. Human neutrophils constitutively express the low-affinity receptors FcγRIIIb (CD16b), FcγRIIIa (CD16a), and FcγRIIa (CD32a)^4–7^. High-affinity FcγRIa (CD64) is also present at low levels and is upregulated in response to G-CSF, during chronic inflammation or sepsis^8,9^. The most abundant receptor on the surface of human neutrophils is FcγRIIIb. It is a glycosylphosphatidylinositol (GPI)-anchored receptor lacking an intracellular domain with specificity for IgG-bound IC and relatively greater affinity to the IgG3 subclass compared to FcγRIIa^10^. The exact mechanisms by which FcγRIIIb crosslinking promotes activation remain elusive^11^. FcγRIIIb signaling has been associated with phagocytosis and the phosphorylation of SYK, which transduces signals leading to ROS production and cytokine expression. Inhibition of SYK following FcγRIIIb crosslinking leads to impaired ROS production^3,12–14^. The lack of intracellular domain suggests that FcγRIIIb signaling relies on co-engagement with other receptors, complicating the identification of its independent role. Others have shown that FcγRIIIb must cooperate with CR3 (CD11b/CD18) or FcγRIIa to mediate ROS production^15–17^. In addition, the seemingly redundant function of FcγRIIIb with other FcγR in neutrophil activation has further complicated dissecting the exact role of FcγRIIIb.

A few studies have reported a rare phenotype of neutrophils lacking FcγRIIIb on their surfaces^4,18–20^. The FcγRIIIb-deficient phenotype may be more frequent in individuals with hematological disorders^21^. One study reported a neonate presenting alloimmune neutropenia while another reported 2 adults with either paroxysmal nocturnal hemoglobinuria or immune neutropenia^18,20^. However, another study reported the absence of FcγRIIIb on the surface of peripheral blood neutrophils collected from an apparently healthy adult individual^4^. This observation may point to a compensated inborn error of immunity, similar to the asymptomatic presentation seen in some cases of myeloperoxidase deficiency^22,23^. To the best of our knowledge, these are the only reports of functional consequences of FcγRIIIb deficiency on neutrophils. The origins of the deficiency have not been described.

Here, we present 2 apparently healthy brothers with FcγRIIIb-deficient neutrophils. In both cases, the absence of FcγRIIIb was caused by missense mutations causing a loss of the start codon in the *FCGR3B* gene. FcγRIIIb**^null^** neutrophils exhibited significantly higher basal expression of FcγRIIIa and FcγRIa. When challenged with opsonized *E. coli* particles, FcγRIIIb**^null^** neutrophils presented significantly higher upregulation of FcγRIa, TLR-4 and αM integrin but reduced L-selectin shedding, actin polymerization, SYK phosphorylation, phagocytosis efficiency, and ROS production. Therefore, this study expands our understanding of FcγRIIIb, the most abundant receptor on FcγRIIIb-expressing neutrophils.

## Materials and methods

### Human samples

This study was approved by the ethics committee of Hospital Materno Infantil from Nuevo Leon, Mexico (DEISC-19 01 24 038). Samples were collected from 10 healthy adult volunteers after informed consent was signed. Briefly, 5 mL of peripheral blood per donor were collected in K_2_-EDTA tubes (BD, cat. 367525) that were kept at room temperature until processing within 30 min of collection. Neutrophils were enriched using a centrifugation gradient using Ficoll-Paque Plus (Cytiva, cat. 17144003). Erythrocytes were lysed using a hypotonic NH_4_Cl buffer, and granulocytes were washed twice with PBS at room temperature. Enrichment purity was assessed by measuring the co-expression of CD11b and CD15 using flow cytometry and was consistently > 98% of double-positive cells. Ficoll-Paque was used to enrich peripheral blood mononuclear cells (PBMC), and residual erythrocytes were lysed using NH_4_Cl.

### DNA sequencing

Genomic DNA was extracted from donor PBMCs and enriched neutrophils using a QIAamp DNA Mini Kit (Qiagen, cat. 51306), following the manufacturer’s protocol. To sequence *FCGR3B* exons, a set of primers was designed to amplify each coding exon of the *FCGR3B* gene using PCR (**Sup. table 1**). Exon sequences were investigated using Sanger technology. Alignments of sequencing results were performed using Geneious prime (Dotmatics, V.2025.0.03).

### Surface immunophenotyping using flow cytometry

Neutrophil surface receptors, Fcγ receptors, and pattern recognition receptors (PRR) were measured by flow cytometry. Briefly, 2×10^5^ cells were incubated on ice with Fc block (Miltenyi Biotech, cat. 130-059-901) for 15 min, then stained with titrated concentrations of CD11b-AF700 (BD, cat. 557918), CD15-BV786 (BD, cat. 563838), CD64-BV650 (BD, cat. 745316), CD32-APC (BD, cat. 559769), CD16b-PE (BD, cat. 550868) and CD16a-AF405 (R&D systems, cat. FAB107512V), or titrated concentrations of TLR-2-AF647 (Biolegend, cat. 309714), TLR-4-APC (Biolegend, cat. 312816), TLR-6-PE (Biolegend, cat. 334708) and CD62L-FITC (BD, cat. 555543) for 30 min at 4 °C in the dark. Monocytes were identified from enriched PBMCs using CD14-PerCP Cy5-5 (Biolegend, cat. 325622), CD16b-PE (BD, cat. 550868) and CD16a-AF405 (R&D systems, cat. FAB107512V).

Fixable viability stain 510 (BD, cat. 564406) was added as a viability marker 15 min before acquisition. Samples were analyzed on a BD FACSCelesta flow cytometer fitted with 405 nm, 488 nm, and 640 nm lasers, operated through the BD® FACSDiva software v.8. Cytometer settings were validated as within the range of manufacturer’s recommendations using CS&T beads (BD®, cat. 642412) prior to each acquisition. Compensation controls were used at each acquisition using the CompBeads anti-mouse Ig, κ/Negative control compensation particle set (BD®, cat. 552843) following manufacturer’s recommendations. At least 20,000 compensated live single events were acquired per sample. Data were analyzed using FlowJo v.10 (BD®).

### Phagocytosis

Phagocytosis assays were performed using opsonized *E. coli* pHrodo bioparticles (Invitrogen, cat. P35361) following the manufacturer’s recommendations, or non-opsonized particles, as stated in figure legends. To produce opsonized bioparticles, 20 μl of heat-inactivated pooled (N=7) sera were added to 20 μl of *E. coli* pHrodo bioparticles and incubated at 37 °C and 100 RPM agitation in a 96-well plate. After 30 min, 1.75 x10^5^ neutrophils were added to the wells and incubated for 30 min at 37 °C and 5% CO_2_ without agitation. The cells were analyzed immediately using a FACSCelesta flow cytometer, as described above. Samples positivity above the background reflected the average proportion of phagocytic cells. A phagocytic index was determined by dividing the MFI of bioparticles-treated cells by the MFI of untreated cells. This index reflects a relative quantity of phagocytosed particles per cell in the population, hence the average relative efficiency of phagocytosis at the cell level.

### Immune complexes (IC) production

To produce IC, *E. coli* DH5-α were cultured in 50 mL of LB medium for 24 h. After incubation, the cells were pelleted at 5,000 rcf, washed twice with a 10% glycerol solution, and aliquoted in 100 μL portions with a concentration 1.44 x10^7^ CFU/mL. Culture aliquots were irradiated with UV light for 3 h and stored at -20 °C until use. Immediately before use, cells were washed with 1 mL of PBS at 16,000 rcf and resuspended in 20 μL of PBS. 20 μL of heat-inactivated pooled serum were added, and the mixture was incubated for 30 min at 37 °C with shaking at 100 rpm. The resulting IC were washed with PBS before use. Some assays were performed with non-opsonized killed *E. coli*, as stated in figure legends. CFU/mL was determined by dilutions plating before irradiation. The ratio IC:neutrophil used for all the experiments using IC was 72:1.

### Actin polymerization

Actin polymerization was investigated using phalloidin staining and measured by flow cytometry. Briefly, 1 x10^6^ neutrophils were stimulated with IC for 0, 15, 30, 60, 180, and 360 sec, followed by immediate fixation with 2% PFA and permeabilization using 0.1% saponin (Sigma, cat. 84510). Phalloidin-AF488 (Invitrogen, cat. A12379) was used for staining, as per manufacturer’s instructions.

### ROS production

ROS production was evaluated by measuring the oxidation of dihydrorhodamine 123 (DHR, Invitrogen, cat. D23806). Briefly, 2 x10^5^ cells/mL in 5 mM DHR were stimulated with either 100 ng/mL of PMA for 15 min at 37 °C or with IC for 30 min at 37 °C. After incubation, the reaction was stopped by a 10 min incubation on ice, cells were centrifuged at 300 rcf for 5 min at 4 °C. DHR conversion was measured immediately by flow cytometry as mentioned above, acquiring >20,000 single events.

### Intracellular pSYK

Briefly, 2 x10^5^ neutrophils were stimulated with IC for 30 min at 37 °C. After incubation, the cells were washed in PBS and centrifuged at 300 rcf for 5 min at 4 °C. Cells were then permeabilized and fixed using the cytofix/cytoperm fixation/permeabilization kit (BD, cat. 554414) and stained with anti-SYK-PE (BD, cat. 558529) as per manufactureŕs instructions and analyzed by flow cytometry as described above. Stimulation indexes were calculated by dividing the MFI of stimulated samples by MFI of non-stimulated samples, these reflect the fold change in pSYK after stimulation.

### Statistical analysis

For each of the 2 available FcγRIIIb**^null^** individuals, >2 technical replicates were measured for every assay, and assays were repeated at least twice on different days. The mean of replicates for each assay performed were plotted on graphs and used for statistical tests. Data distributions were evaluated using Shapiro-Wilk tests. Comparisons between 2 groups were performed using unpaired student t-test. All statistical tests were performed using Prism V.9 software (GraphPad). P-values < 0.05 were considered significant.

## Results

### Mutations in exon 2 of the *FCGR3B* gene caused the loss of FcγRIIIb translation

We serendipitously identified 2 individuals lacking FcγRIIIb expression on peripheral blood neutrophils using flow cytometry **(Sup. Fig. 1)**. To investigate if the absence of FcγRIIIb had a genetic origin, we obtained DNA from neutrophils and PBMCs from these donors and sequenced the 5 coding exons of the *FCGR3B* gene using Sanger sequencing. Consistently and irrespective of the source of DNA, exons showed >95% identity with the *FCGR3B* reference sequence (RefSeq ID 2215, **Sup. Table 2**). However, for both donors, exon 2 exhibited only 52% identity with the reference sequence. Nucleotide sequence analysis confirmed an ATG to CCC mutation leading to a methionine to proline substitution, leading to the absence of the original start codon and an altered open reading frame that included multiple stop codons **(Sup. Fig. 2 and 3**). Alignment of the *FCGR3B* exon sequences from the 2 FcγRIIIb**^null^** individuals showed the same mutations in exon 2 in both individuals **(Sup. Fig. 4)**. Therefore, a mutation causing the loss of a start codon in exon 2 of *FCGR3B* prevented the translation of FcγRIIIb. To the best of our knowledge, this is the first report identifying a genetic cause for the FcγRIIIb-deficient phenotype.

### Fc**γ**RIIIb^null^ neutrophils phagocytosed less and exhibited defects in actin polymerization

Given the pivotal role of FcγRIIIb in binding IC and activating neutrophils, we investigated if lacking FcγRIIIb altered neutrophil antimicrobial functions. Challenge with IC promoted phagocytosis in >95% neutrophils, irrespective of the presence of FcγRIIIb, as evidenced by the proportion of cells exhibiting fluorescence above the threshold for positivity, indicative of oxidation of the pHrodo bioparticles **(Fig. 1A)**. However, comparing phagocytic indexes indicated that FcγRIIIb**^null^**neutrophil phagocytosed on average significantly less IC, compared to FcγRIIIb-expressing neutrophils (p = 0.0007, **Fig. 1B**). Phagocytosis depends on actin remodeling^24–26^. Thus, we compared actin polymerization upon IC stimulation at various time points immediately post-IC challenge. FcγRIIIb**^null^**neutrophils had significantly lower actin polymerization at 15 s, 30 s and 60 s (p = 0.024, p = 0.0034, and p = 0.001, respectively, **Fig. 1C**) compared to FcγRIIIb-expressing neutrophils. These results suggest a critical role of FcγRIIIb in regulating early actin cytoskeleton remodeling, and phagocytosis upon crosslinking with IC.

**Figure 1.**
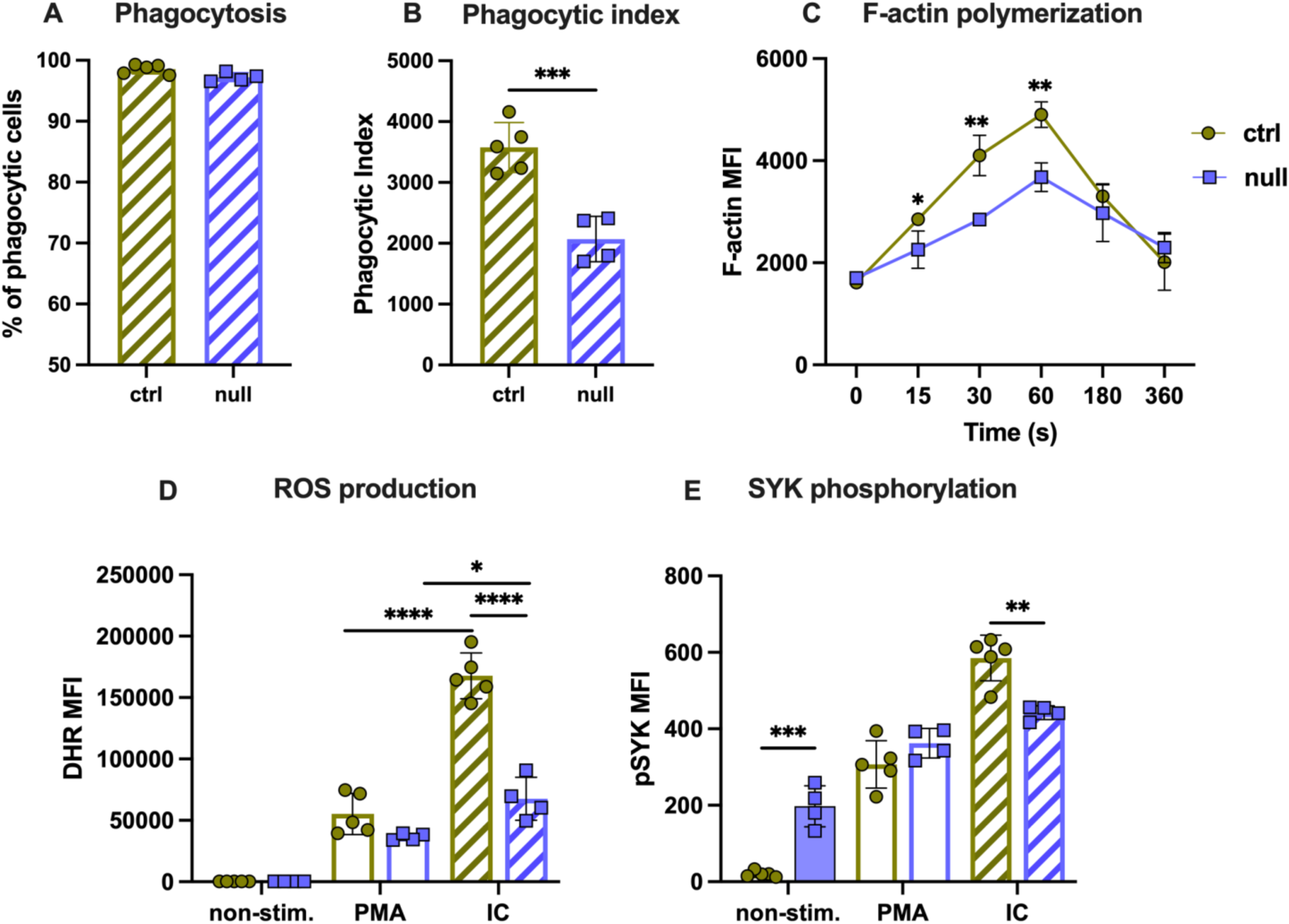
Phagocytosis and ROS production are impaired in FcγRIIIb^null^ neutrophils. (A) Proportion of phagocytosing cells per sample. (B) Phagocytic index, calculated by dividing the MFI of bioparticles-treated cells by the MFI of untreated cells. This index reflects a relative quantity of phagocytosed particles per cell in the population. (C) F-actin polymerization following IC stimulation, (N = 4 biological samples for the control group and 2 for the null group, with >1 technical replicate performed on different days). (D) ROS production. (E) Median fluorescence intensity (MFI) of pSYK following PMA and IC stimulation. Peripheral blood FcγRIIIb-expressing neutrophils (ctrl, green circles). Peripheral blood FcγRIIIb^null^ neutrophils (null, purple squares). IC: stimulation with immune complexes. N = 5 biological replicates for the control group and 2 biological replicates for the null group. Measurements performed in duplicates, performed on different days. ⍰=p<0.05, ⍰⍰=p<0.01, ⍰⍰⍰ = 0.001, ⍰⍰⍰⍰ p<0.0001.

### FcγRIIIb^null^ neutrophils exhibited decreased Fc**γ**R-mediated ROS production

We then investigated the capacity of FcγRIIIb**^null^** neutrophils to produce ROS. PMA stimulation led to similar levels of ROS, irrespective of the expression of FcγRIIIb (**Fig. 1D**). This was expected as PKC activation with PMA bypasses activation through FcγR crosslinking. This suggested that part of the ROS production pathway, starting from PKC activation, was not affected by the absence of FcγRIIIb. IC stimulation was significantly more efficient than PMA at eliciting ROS production (**Fig. 1D**). However, we measured significantly lower ROS produced from FcγRIIIb**^null^** neutrophils compared to FcγRIIIb-expressing neutrophils (p = 0.00007, **Fig. 1D**). These results suggest that FcγRIIIb has a significant role in connecting IgG crosslinking to PKC activation and possibly other routes that culminate in ROS production.

### Fc**γ**RIIIb deficiency increased basal SYK phosphorylation levels

Previous reports have shown that FcγR-mediated ROS production depends on SYK phosphorylation, an early step in FcγR-mediated intracellular cascade^12,27^. To study if FcγRIIIb deficiency impacted SYK-mediated signaling, we investigated intracellular SYK phosphorylation (pSYK). In the absence of stimulation, FcγRIIIb**^null^** neutrophils had significantly higher levels of phosphorylated SYK compared to FcγRIIIb-expressing neutrophils (p = 0.00014, **Fig. 1E**). It is unclear whether this increase in pSYK correlates with significantly more SYK in FcγRIIIb-deficient neutrophils. PMA stimulation resulted in increased pSYK irrespective of FcγRIIIb presence (**Fig. 1E**). These results parallel the PMA-mediated ROS production that was similar irrespective of the presence of FcγRIIIb **(Fig. 1D)**. While PMA stimulation directly activates PKC, this stimulation also causes the phosphorylation of SYK by ROS^28,29^. Therefore, the increase in pSYK in FcγRIIIb-deficient neutrophils may result from PKC-mediated ROS production. IC stimulation also resulted in significantly higher pSYK overall (p < 0.0001 in both FcγRIIIb**^null^**neutrophils and FcγRIIIb-expressing neutrophils, **Fig. 1E**). However, there was significantly less pSYK in IC-stimulated FcγRIIIb**^null^**neutrophils compared to FcγRIIIb-expressing neutrophils (p = 0.0025, **Fig. 1E**). This parallels previous observation in FcγRIII-deficient mouse macrophages upon IgG-crosslinking^30^. While stimulation increased pSYK irrespective of the presence of FcγRIIIb, the upregulation was incremental in FcγRIIIb**^null^**neutrophils, as the basal pSYK levels were significantly elevated compared to FcγRIIIb-expressing neutrophils (mean stimulation index in IC-stimulated groups: 2.39 vs. 26.6, respectively, p = 0.0002). This is consistent with FcγRIIIb crosslinking involved in SYK phosphorylation^12^. These results suggest that the impaired ROS production observed in FcγRIIIb**^null^** neutrophils may originate at least in part from alterations in SYK activation.

### Fc**γ**RIIIb^null^ neutrophils upregulated Fc**γ**RIIIa and Fc**γ**RIa in basal and stimulated states

We investigated whether functional impairments in the absence of FcγRIIIb may be associated with altered expression of other receptors on neutrophil surfaces. FcγRIIIb-expressing neutrophils showed high levels of surface FcγRIIIb in the absence of stimulation. Upon IC stimulation, FcγRIIIb expression was reduced 5-fold (p = 0.0004, **Fig. 2A**). This was consistent with previous studies showing that crosslinking of FcγRIIIb led to a reduction in its surface expression, which may be caused by internalization during phagocytosis or metalloproteinase-mediated cleavage^31,32^. The expression of FcγRIIa was similar between FcγRIIIb**^null^** neutrophils and FcγRIIIb-expressing neutrophils in the absence of stimulation (**Fig. 2B**), suggesting the regulation of this receptor may be independent of FcγRIIIb. In agreement with this, FcγRIIa expression was similarly reduced upon IC-challenge in both FcγRIIIb**^null^** neutrophils (p < 0.0001, **Fig. 2B**) and FcγRIIIb-expressing neutrophils (p < 0.0001, **Fig. 2B**). In contrast, FcγRIIIa expression was significantly higher in FcγRIIIb**^null^** neutrophils compared to FcγRIIIb-expressing neutrophils in the absence of stimulation (p = 0.00004, **Fig. 2C**). Upon IC stimulation, the expression of FcγRIIIa was significantly reduced on FcγRIIIb**^null^** neutrophils only (p < 0.0001, **Fig. 2C**). This suggests a possible involvement in phagocytosis or activation-mediated-cleavage. FcγRIa was also significantly elevated in FcγRIIIb**^null^** neutrophils compared to FcγRIIIb-expressing neutrophils in the absence of stimulation (p < 0.00001, **Fig. 2D**). Upon IC challenge, FcγRIa was upregulated in FcγRIIIb-expressing neutrophils (**Fig. 2D**), as expected^33^. FcγRIIIb**^null^** neutrophils also exhibited increased FcγRIa in this condition, although the increase was significantly higher compared to FcγRIIIb-expressing neutrophils (p = 0.005, **Fig. 2D**). FcγRIa contributes to phagocytosis, although to a lesser extent compared to FcγRIIa^34^. As FcγRIIa was not upregulated in FcγRIIIb**^null^**neutrophils, it is possible that the increase in FcγRIa correlates with an attempt at compensating for the absence of FcγRIIIb to maintain antimicrobial functions. We investigated possible systemic regulations in peripheral blood. There were similar proportions of monocyte populations irrespective of FcγRIIIb status (**Sup.** Fig. 5A-B). Non-classical monocytes did not express FcγRIIIb (data not shown) and in the relative expression of FcγRIIIa was similar irrespective of FcγRIIIb status (**Sup. Fig. 5C-D**).

**Figure 2.**
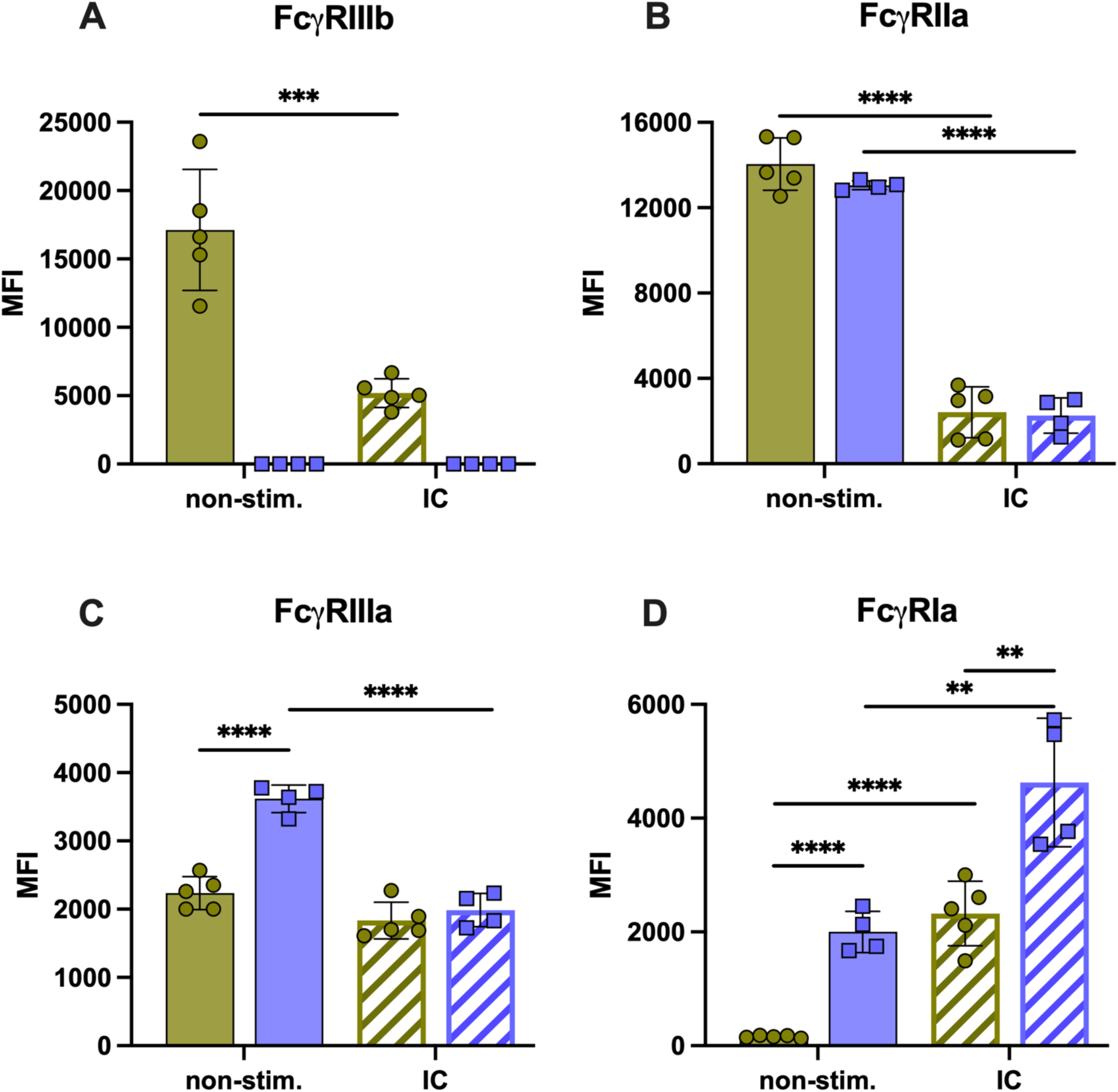
FcγRIIIa and FcγRIa are significantly upregulated on the surface of peripheral blood FcγRIIIb^null^ neutrophils. (A) Median fluorescence intensity (MFI) of surface Fcγ receptors in the absence of stimulation and following stimulation with immune complexes (IC) for FcγRIIIb, (B) FcγRIIa, (C) FcγRIIIa, and (D) FcγRIa. Peripheral blood FcγRIIIb-expressing neutrophils (ctrl, green circles). Peripheral blood FcγRIIIb^null^ neutrophils (null, purple squares). N = 5 biological samples for the control group (ctrl) and 2 biological replicates for the null group (null), with 2 technical replicates each, performed on different days. ⍰⍰=p<0.01, ⍰⍰⍰⍰ p<0.0001.

### Fc**γ**RIIIb^null^ neutrophils upregulated TLR-4 upon stimulation

Having evidenced regulations of other FcγR in FcγRIIIb**^null^**, we wondered if additional receptors relevant in direct microbial recognition would be regulated in this context. TLR-2 recognizes diacylated lipoproteins from bacteria and fungi, resulting in activation of signaling pathways leading to phagocytosis and cytokine production^35,36^. In the absence of stimulation, TLR-2 had a similar expression on neutrophil surfaces, irrespective of FcγRIIIb (**Fig. 3A**). Following challenge with UV-killed *E. coli,* TLR-2 was similarly significantly upregulated on neutrophil surfaces (**Fig. 3A**). TLR-2 was also significantly upregulated after IC stimulation, irrespective of FcγRIIIb (**Fig. 3A**). This overall suggested the regulation of TLR-2 is independent of FcγRIIIb. TLR-6 recognizes lipoproteins present in gram-positive bacteria. There was no difference in TLR-6 expression between groups, irrespective of FcγRIIIb, or stimulation (**Fig. 3B**). This is in line with *E. coli* being a gram-negative bacteria not activating TLR-6 and suggests the regulation of TLR-6 is independent of FcγRIIIb. TLR-4 is the major receptor for lipopolysaccharides (LPS) present on gram-negative bacteria^37^. In the absence of stimulation, TLR-4 was present at similar levels irrespective of FcγRIIIb (**Fig. 3C**). Surprisingly, upon challenge with non-opsonized UV-killed *E. coli* there was a significant upregulation of TLR-4 on the surface of FcγRIIIb**^null^** neutrophils only (p = 0.02, **Fig. 3C**), and this upregulation was also measured after IC stimulation (p = 0.004, **Fig. 3C**). We argue that LPS from *E. coli* may protrude from IC, activating TLR-4 to similar levels as non-opsonized *E. coli*. Stimulation with non-opsonized *E. coli* particles prompted similar levels of phagocytosis irrespective of the presence of FcγRIIIb (**Fig. 3D**). The phagocytosis indexes using *E. coli* particles also suggested similar efficiency of phagocytosis irrespective of the presence FcγRIIIb (**Fig. 3E**). This contrasted with the lower phagocytosis efficiency of FcγRIIIb**^null^** neutrophils using opsonized pHrodo *E. coli* particles (**Fig. 1B**). ROS production was also increased upon non-opsonized *E. coli* stimulation irrespective of the presence FcγRIIIb (p = 0.0007, **Fig. 3F**), again contrasting with the impairment seen in FcγRIIIb**^null^**neutrophils stimulated with IC. Of note, when neutrophils were stimulated through TLR-4 in the absence of FcγR crosslinking, the relative increase in ROS and phagocytosis efficiency were not as high as when neutrophils were stimulated with IC, that stimulate both TLR-4 and FcγR. Studies on the relative strength of neutrophil activation depending on stimulation through TLR-4 versus through FcγR are limited. However, other have shown that in the absence of TLR-4, neutrophils do not get activated by IC. TLR-4 co-localized in lipid rafts with FcγRs upon FcγR engagement, and TLR-4 was necessary for ITAM phosphorylation downstream of most FcγR^38,39^. This suggested a crosstalk and regulation of FcγR by TLR-4^38^. Here, in the absence of FcγRIIIb, neutrophils significantly upregulate TLR-4 when challenged with gram-negative bacteria or IC derived from gram-negative bacteria. This significant upregulation could originate from the lack of a negative feedback that may happen after crosslinking FcγRIIIb in FcγRIIIb-expressing neutrophils. This also suggests a possible novel direct regulation of TLR-4 functions by FcγRIIIb.

**Figure 3.**
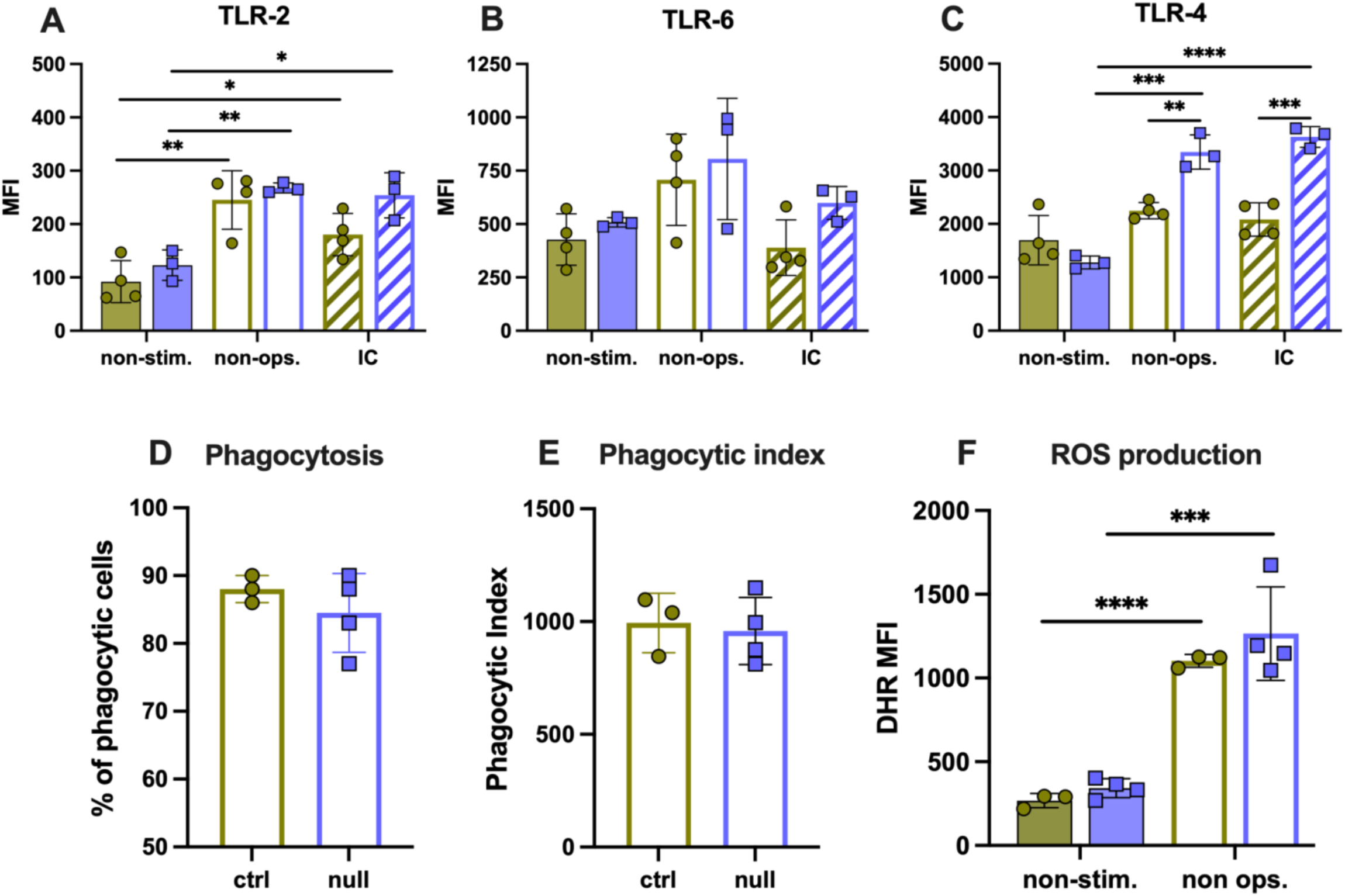
TLR-4 is significantly upregulated on the surface of FcγRIIIb^null^ neutrophils upon stimulation with *E. coli*. Median fluorescence intensity (MFI) of (A) TLR-2, (B) TLR-6, and (C) TLR-4 in the absence of stimulation (non-stim), and following stimulation with UV-killed *E. coli* immune complexes (IC). N = 4 biological samples for the control group (ctrl) and 1 for the null group (null), with 3 technical replicates each, performed on different days. (D) Percentage of phagocytosing cells per sample. (E) Phagocytic index calculated by dividing the MFI of IC-treated cells by the MFI of untreated cells. This index reflects a relative quantity of phagocytosed particles per cell in the population. (F) ROS production following stimulation with non-opsonized UV-killed *E. coli.* N = 3 biological samples for the control group (ctrl) and 2 for null group (null) with 2 technical replicates each, performed on different days. Peripheral blood FcγRIIIb-expressing neutrophils (ctrl, green circles). Peripheral blood FcγRIIIb^null^ neutrophils (null, purple squares). Non-ops: stimulation with non-opsonized UV-killed *E. coli*, IC: stimulation with immune complexes. ⍰⍰=p<0.01, ⍰⍰⍰ = 0.001.

### Adhesion molecules are differentially regulated in Fc**γ**RIIIb^null^ neutrophils upon stimulation

We finally investigated receptors involved in neutrophil trafficking. In the absence of stimulation, the levels of αM integrin (CD11b) and L-selectin (CD62L) were similar on neutrophil surfaces irrespective of FcγRIIIb (**Fig. 4**). Upon stimulation with either non-opsonized *E. coli* or IC, αM integrin was significantly increased on the surface of neutrophils (**Fig. 4A**). The increase was more significant in FcγRIIIb**^null^**neutrophils compared to FcγRIIIb-expressing neutrophils (**Fig. 4A**). The significantly higher expression of αM integrin in FcγRIIIb**^null^** neutrophils may improve complement-mediated phagocytosis, as IC phagocytosis was impaired (**Fig. 1B-C**). While neutrophils shed L-selectin upon activation, as expected, FcγRIIIb**^null^** neutrophils exhibited significantly less shedding compared to FcγRIIIb-expressing neutrophils, irrespective of the type of stimulus (p = 0.022 and 0.0021 for each type of stimulation, **Fig. 4B**). Overall, these different regulations of adhesion molecules suggest possible altered neutrophil-endothelial interactions, i.e. rolling, arrest, and crawling in the absence of FcγRIIIb^40^.

**Figure 4.**
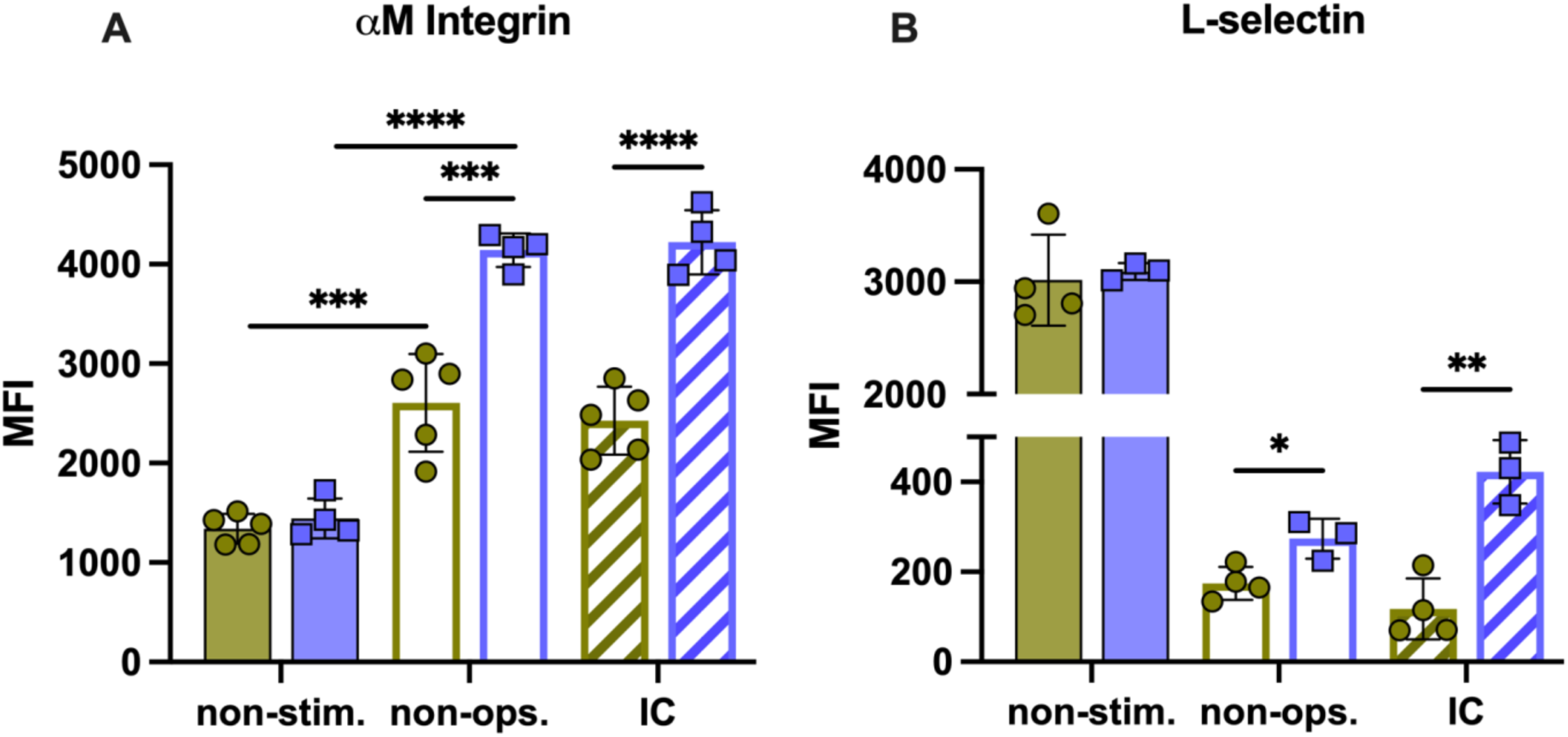
CD11b and CD62L regulated in FcγRIIIb^null^ neutrophils. Median fluorescence intensity (MFI) of (A) αM integrin (CD11b) and (B) L-selectin (CD62L). Peripheral blood FcγRIIIb-expressing neutrophils (ctrl, green circles). Peripheral blood FcγRIIIb^null^ neutrophils (null, purple squares). Non-stim.: non-stimulated cells, Non-ops: stimulation with non-opsonized bacteria UV-killed *E. coli*, IC: stimulation with immune complexes. N = 5 biological sample for the control group and 2 samples for the null group, with 2 technical replicates each, performed on different days. ⍰=p<0.05, ⍰⍰=p<0.01, ⍰⍰⍰ = 0.001 ⍰⍰⍰⍰ p<0.0001.

## Discussion

Here, we report 2 apparently healthy brothers presenting FcγRIIIb deficiency on neutrophils. Missense mutations in exon 2 of the *FCGR3B* gene affected the start codon, leading to loss of translation of the FcγRIIIb protein. Earlier studies using southern blots or allele-specific PCR have proposed that complete gene deletion was the cause of FcγRIIIb-deficiency^41,42^. As these techniques rely on the presence of specific sequences targeted by primers, mutations in the sequences could have hampered the binding of primers. To the best of our knowledge, this is the first report to provide exon sequence data from FcγRIIIb**^null^** individuals, identifying a genetic cause of the FcγRIIIb**^null^**phenotype.

We describe various impairments of the neutrophil response that correlated with the absence of FcγRIIIb. A previous study described that FcγRIIIb-deficient neutrophils maintain normal phagocytosis^4^. However, that study reported the percentage of phagocytosing cells only. While we also report unaffected proportions of phagocytosing cells, the phagocytic index revealed an overall significant impairment in phagocytosis efficiency in FcγRIIIb**^null^** neutrophils. Others have shown that FcγRIIa crosslinking was more efficient than FcγRIIIb at triggering phagocytosis, using selective agonists^6^. Here, the absence of FcγRIIIb did not impact FcγRIIa expression levels. Altogether, our data suggest that FcγRIIIb has a pivotal role during phagocytosis as its absence leads to reduced phagocytic efficiency. This could also be explained by defects in actin polymerization following FcγR stimulation in FcγRIIIb**^null^** neutrophils^24^.

ROS production was significantly impaired in FcγRIIIb**^null^** neutrophils. Others have shown that FcγRIIa crosslinking results in lower ROS production compared to FcγRIIIb crosslinking^12^. Therefore, the ROS produced in FcγRIIIb**^null^** neutrophils upon IC challenge could be triggered through FcγRIIa, as this remained stably expressed in FcγRIIIb**^null^** neutrophils. The reduced ROS production in FcγRIIIb**^null^**neutrophils evidenced the crucial role of FcγRIIIb in ROS generation. Although FcγRIIa can trigger ROS, it is not sufficient to fully restore the ROS response in FcγRIIIb**^null^** neutrophils.

SYK phosphorylation is crucial in early neutrophil activation events triggered by FcγRs crosslinking, culminating in phagocytosis, ROS production, and cytokine release^3,13,43,44^. Inhibition of SYK decreased FcγRIIIb- and FcγRIIa-mediated ROS production^12^. Increased SYK levels may lead to increased basal phosphorylation, even in the absence of stimulation. This could be due to the activity of other kinases, autophosphorylation, or mutations in the COOH-terminal region of SYK^45^. This may correlate with a higher concentration of total SYK in these individuals. Lower levels of pSYK in FcγRIIIb**^null^** neutrophils compared to FcγRIIIb-expressing neutrophils upon IC challenge indicate that FcγRIIIb crosslinking may be involved in SYK phosphorylation. To the best of our knowledge, the total SYK levels in FcγRIIIb-deficient neutrophils have not been measured and compared to FcγRIIIb-expressing neutrophils. Lower pSYK in FcγRIIIb**^null^** neutrophils upon IC crosslinking may result from a diminished capacity to activate downstream signaling molecules in the absence of FcγRIIIb, thus highlighting the critical role of FcγRIIIb in activating pSYK and subsequent ROS production.

Others have suggested that FcγRIIa and FcγRIIIa may compensate for the absence of FcγRIIIb, as these receptors were capable of initiating phagocytosis and cell activation in FcγRIIIb**^null^** individuals^4^. FcγRIIIa is a high-affinity FcγR, initially thought to be restricted to NK cells and monocytes^46,47^. FcγRIIIa was identified on neutrophil surfaces, where it contributes to neutrophil activation and phagocytosis upon crosslinking with antibody-opsonized beads. The same authors reported that blocking FcγRIIIa/b in FcγRIIIb-expressing neutrophils resulted in significantly reduced phagocytosis compared to FcγRIIIb**^null^**neutrophils, suggesting that FcγRIIIb plays a more significant role in mediating phagocytosis than FcγRIIIa^4^. Here, FcγRIIIb**^null^** neutrophils exhibited significantly higher levels of FcγRIIIa compared to FcγRIIIb-expressing controls in the absence of stimulation. Interestingly, upon IC stimulation, FcγRIIIa was downregulated only in FcγRIIIb**^null^** neutrophils, suggesting its potential internalization during phagocytosis, as reported in FcγRIIIb-deficient neutrophils^4^.

FcγRIa is expressed at low levels on human neutrophils but is upregulated during infections, stimulation with proinflammatory cytokines, or G-CSF^8,9,48^. The relevance of increased FcγRIa in the absence of FcγRIIIb is unclear. Indeed, IFN-γ-induced FcγRIa was unable to mediate phagocytosis of IgG-opsonized fluorescent latex particles^6^. The absence of FcγRIa downregulation post-IC challenge in FcγRIIIb**^null^**neutrophils, suggests that FcγRIa is not involved in phagocytosis, in agreement with earlier results^6^. Despite upregulations of FcγRIa and FcγRIIIa in the absence of FcγRIIIb, phagocytosis efficiency and ROS production upon IC challenge were impaired. This is in line with these receptors being secondary to FcγRIIIb and FcγRIIa in the clearance of opsonized bacteria^6,49,50^. It was intriguing to measure consistent FcγRIIa in the absence of FcγRIIIb, given the upregulation of other FcγRs and the usual high expression of both receptors on neutrophil surfaces. This may contrast with the earlier hypothesis that FcγRIIIb signaled through dimerizing with FcγRIIa^16,51^.

TLRs activate neutrophils through PAMPs, thus mediating key functions^52^. In an earlier study in mice, FcγRIII^-/-^ neutrophils could produce cytokines upon LPS or IgG stimulation. However, neutrophils lacking TLR-4 produced less cytokines upon IgG stimulation, suggesting a potential crosstalk between FcγRIII and TLR-4^38^. Here, we found similar phagocytosis and ROS production after non-opsonized *E. coli* challenge with FcγRIIIb**^null^**neutrophils, that correlated with higher TLR-4 in these cells. Therefore, in these individuals, TLR-mediated functions may be enhanced, and TLR-4 may be regulated by FcγRIIIb. Additional research should investigate further TLR-mediated functions in FcγRIIIb**^null^** neutrophils. Specifically, TLRs play a crucial role in regulating cytokine production in response to bacterial recognition^35,38^.

Here, we found that αM integrin upregulation upon stimulation was significantly higher in FcγRIIIb**^null^**neutrophils. This correlated with lower downregulation of L-selectin in the same conditions, altogether describing altered regulations of molecules involved in neutrophil rolling and adhesion. This also suggests that the upregulation of αM integrin observed in FcγRIIIb**^null^** neutrophils may be associated with an enhanced ability to recognize and eliminate complement-opsonized pathogens. It will be relevant to investigate if FcγRIIIb plays a role in neutrophil transendothelial migration, which would not be surprising considering also the defect in actin polymerization in FcγRIIIb**^null^** neutrophils.

In conclusion, we characterized the genetic origin of FcγRIIIb deficiency on the surface of neutrophils in 2 brothers. Various bactericidal functions were impaired in FcγRIIIb**^null^**neutrophils, including actin polymerization, phagocytosis, and ROS production following challenge with IC. The absence of FcγRIIIb significantly altered the landscape of receptors on the neutrophil surface with distinct differences between basal conditions and after IC stimulation. This work expands our understanding of the role of FcγRIIIb in neutrophils, including a previously unrecognized role in regulating the expression of TLR-4, other FcγRs, and adhesion molecules.

## Data Availability

All data produced in the present study are available upon reasonable request to the authors

## Acknowledgments

We thank Centro de Biotecnología FEMSA from Tecnológico de Monterrey for equipment maintenance. We acknowledge the financial support received by Secretaría de Ciencia, Humanidades, Tecnología e Innovación (Secihti, scholarships 1007842, 1105685, and 657487).

## Author contribution statement

JACC designed and performed experiments, analyzed data, produced figures, wrote and edited the manuscript. JACP, performed phagocytosis experiments, analyzed data, wrote and edited the manuscript. ALA, performed ROS production experiments, analyzed data, wrote and edited the manuscript. ABSA, designed and performed actin polymerization experiments, analyzed data, wrote and edited the manuscript. MS, analyzed data, wrote and edited the manuscript. MEGB, conceived the study, secured funding, analyzed data, wrote and edited the manuscript. All authors approved the final version of the manuscript.

---

**Supplementary table 1.**
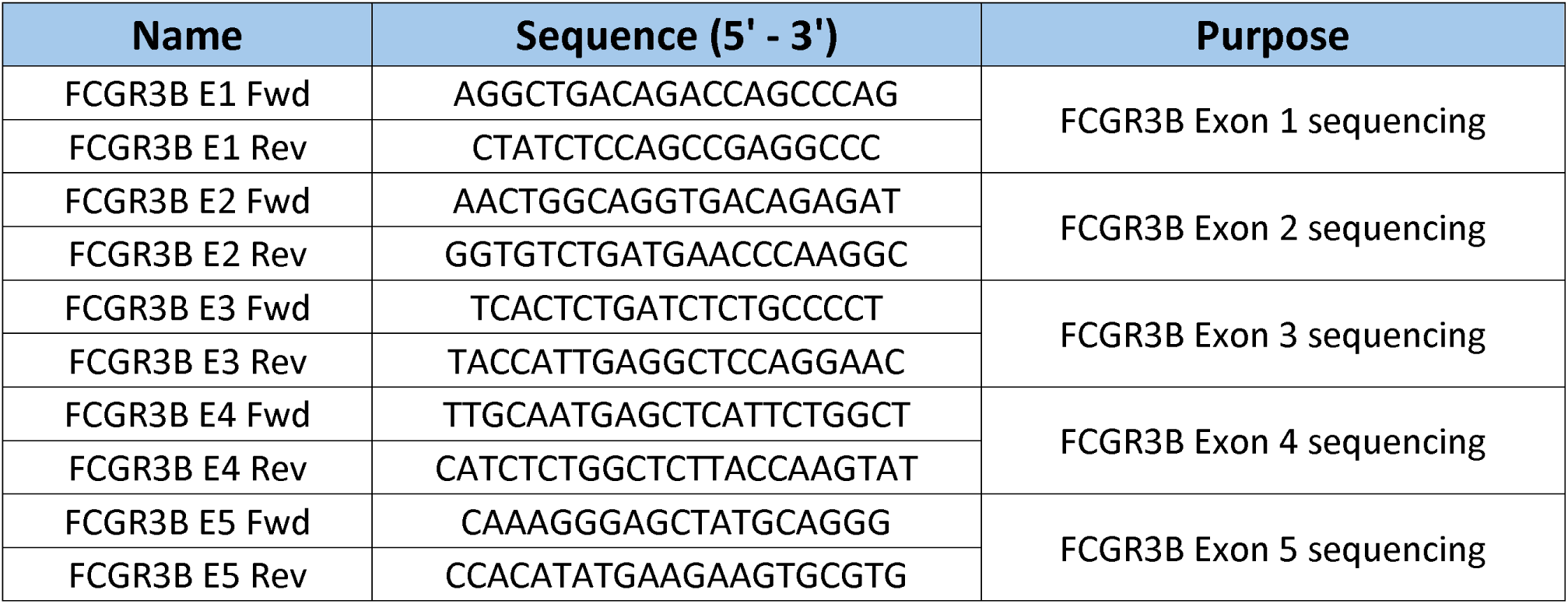
Primers sequences.

**Supplementary table 2.** FCGR3B sequence identity between the consensus reference sequence (Genbank ID 2215) and sequences from donors of neutrophils lacking FcγRIIIb expression.

| Sequence name | % Identity to refseq (Genbank ID 2215) |  |
| --- | --- | --- |
|  | Individual 1 | Individual 2 |
| Exon 1 | 96.16% | 92.50% |
| Exon 2 | 51.18% | 52.83% |
| Exon 3 | 100% | 100% |
| Exon 4 | 100% | 100% |
| Exon 5 | 96.53% | 94.98% |

## Figure legends

**Supplemental Figure 1.**
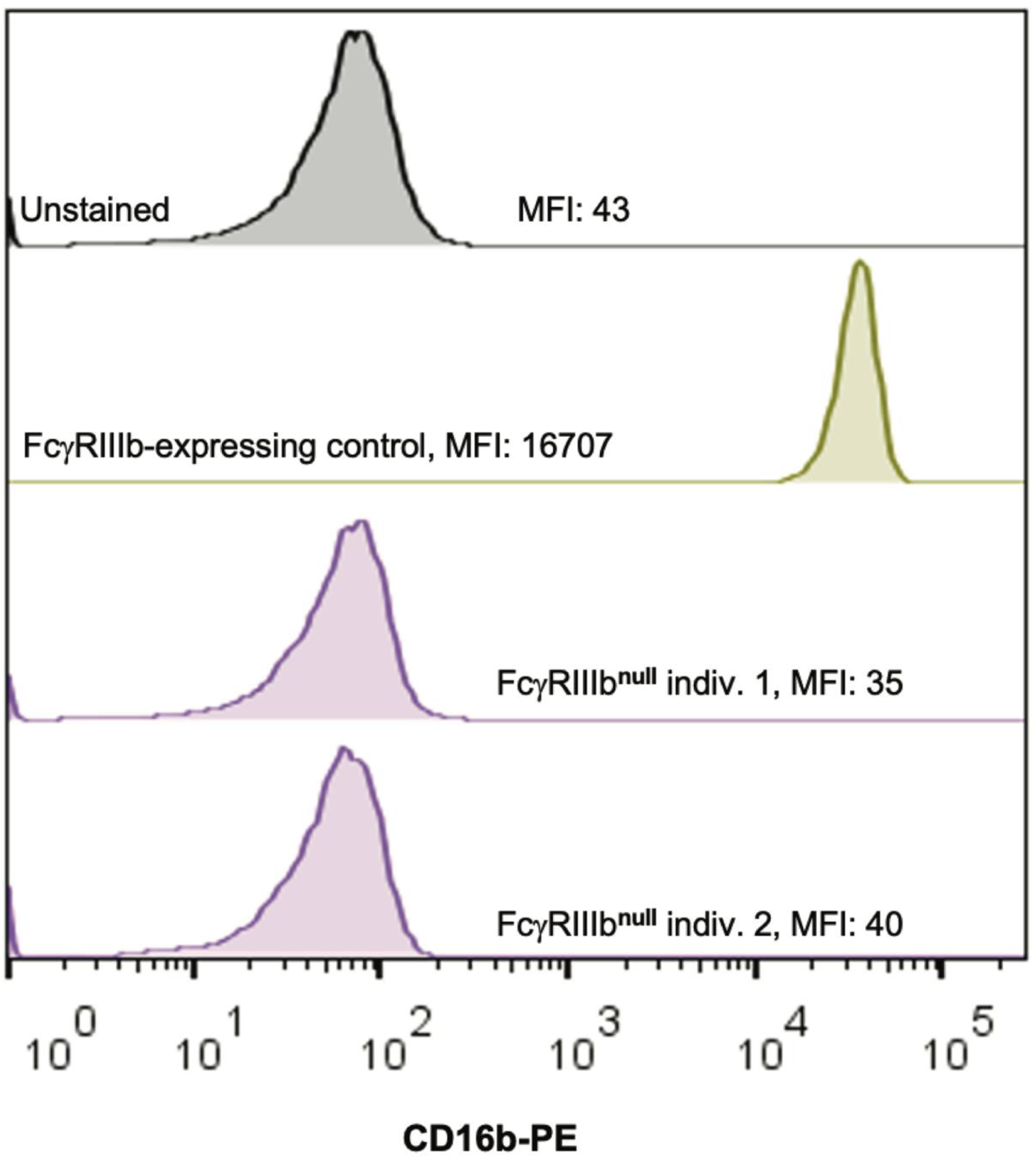
Identification of FcγRIIIb^null^ individuals. Representative histograms of FcγRIIIb-expressing and FcγRIIIb-lacking neutrophils.

**Supplemental Figure 2.**
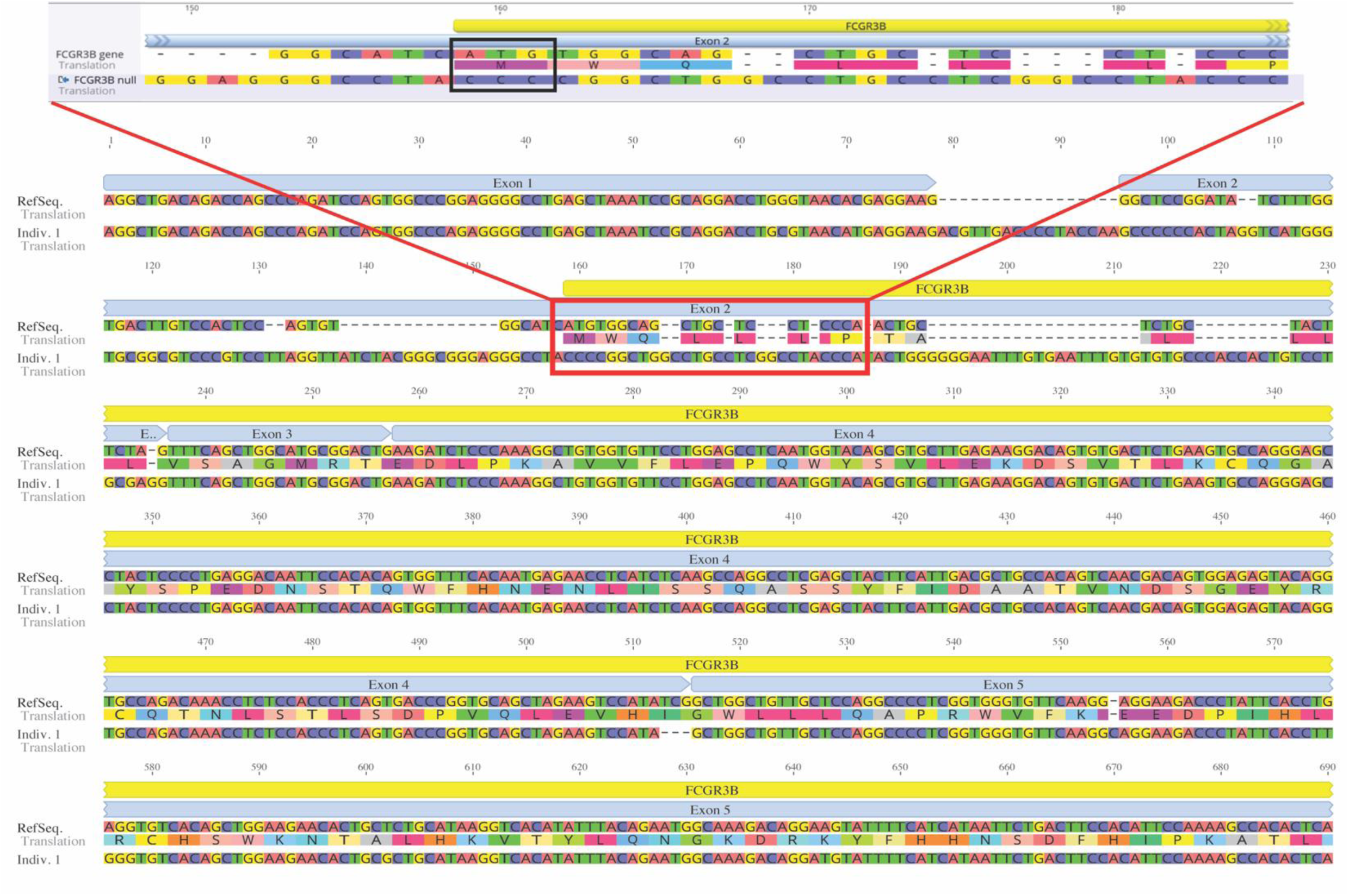
DNA alignment of FCGR3B gene in individual 1 lacking FcγRIIIb expression. The highlighted red rectangle represents the start of protein translation, evidencing mutations on the start codon leading to translation loss. The black rectangle indicates mutations on the start codon, leading to translation loss.

**Supplemental Figure 3.**
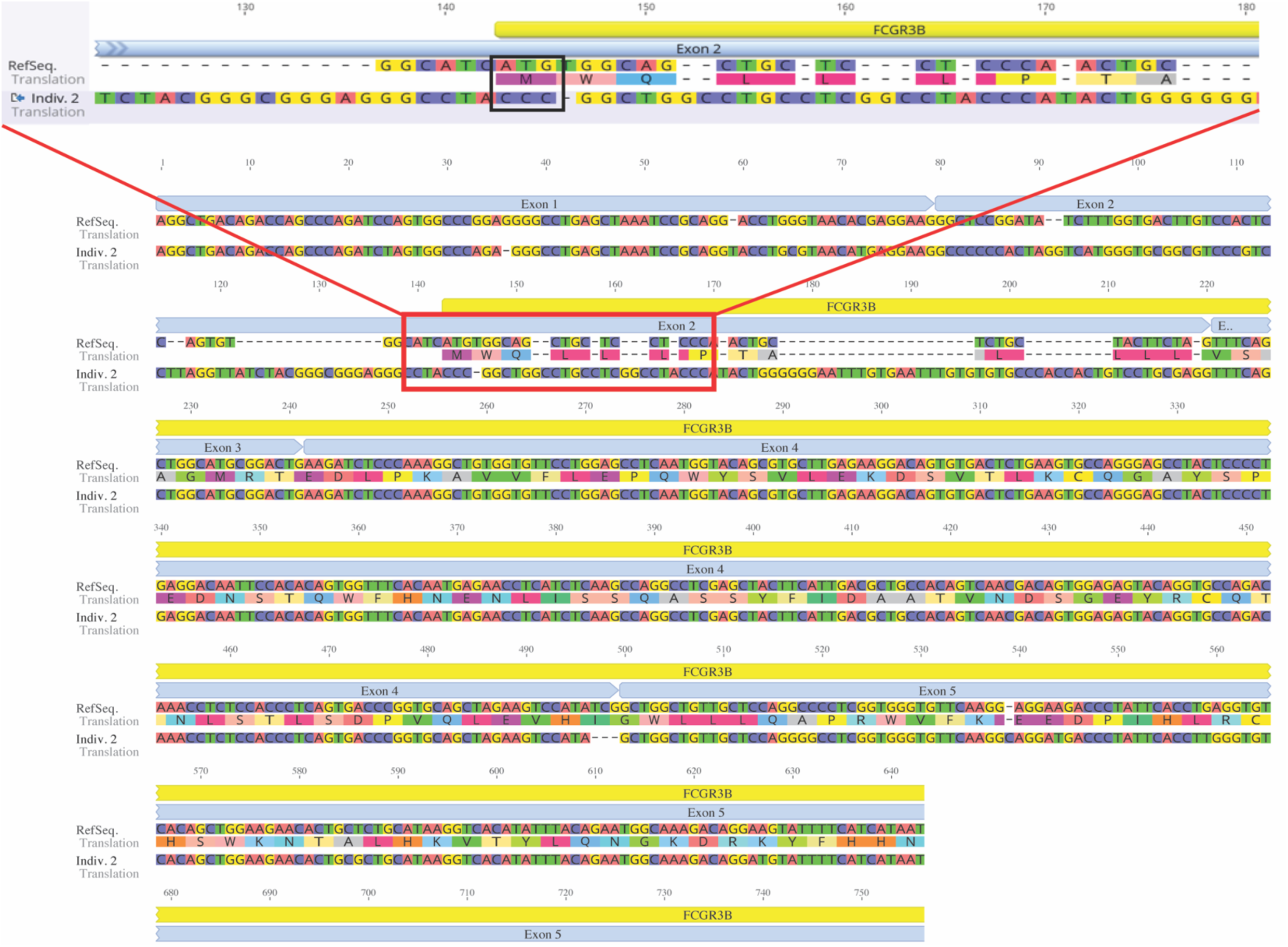
DNA alignment of FCGR3B gene in individual 2 lacking FcγRIIIb expression. The highlighted red rectangle represents the original start of protein translation. The black rectangle indicates the mutations affecting the start codon, leading to translation loss.

**Supplemental Figure 4.**
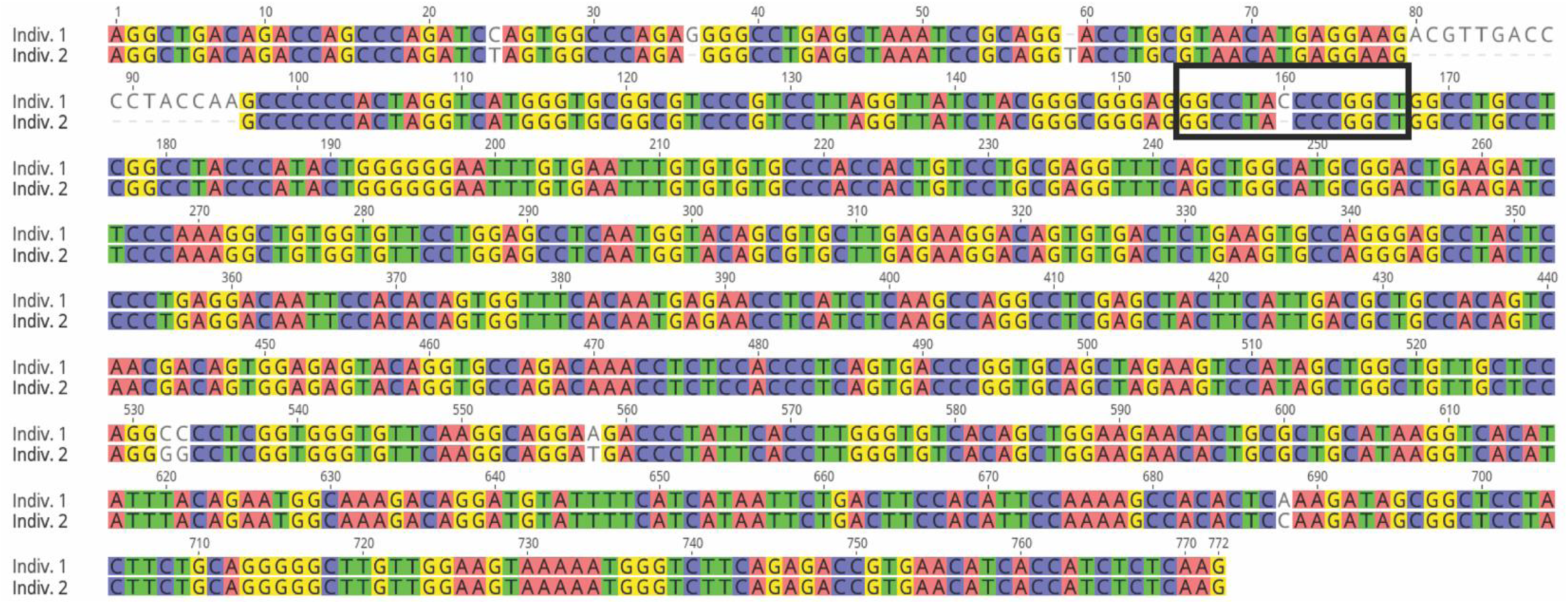
DNA alignment of FCGR3B between FcγRIIIb^null^ individuals. The black rectangle indicates the mutations affecting the start codon, leading to translation loss.

**Supplemental Figure 5.**
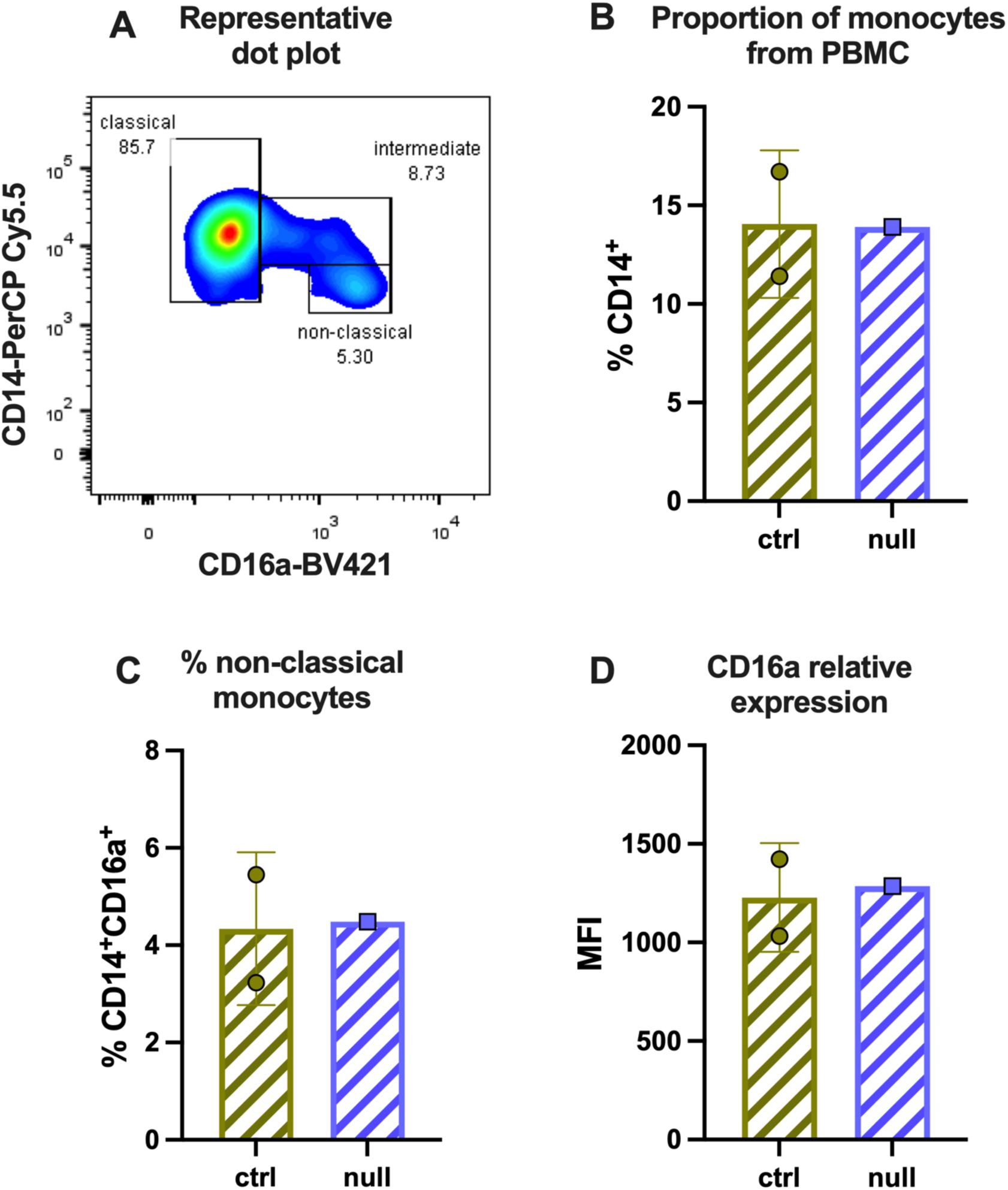
Monocytes proportions in FcγRIIIb^null^ individuals. (A) Representative gating in CD14 vs. CD16a density plot of PBMC monocytes. (B) Proportions of monocytes in FcγRIIIb-expressing and FcγRIIIb^null^ individuals (C) Proportions of non-classical FcγRIIIa-expressing monocytes in the peripheral blood from FcγRIIIb-expressing and FcγRIIIb^null^ individuals. (D) MFI of FcγRIIIa present on the surface of non-classical monocytes. Peripheral blood FcγRIIIb-expressing neutrophils (ctrl, green circles). Peripheral blood FcγRIIIb^null^ neutrophils (null, purple squares).

## Notes

**Conflicts of Interest:** None.

### Competing Interest Statement

The authors have declared no competing interest.

### Funding Statement

We thank Centro de Biotecnologia FEMSA from Tecnologico de Monterrey for equipment maintenance. We acknowledge the financial support received by Secretaria de Ciencia, Humanidades, Tecnologia e Innovacion (Secihti, scholarships 1007842, 1105685, and 657487).

### Author Declarations

This study was approved by the ethics committee of Hospital Materno Infantil from Nuevo Leon, Mexico (DEISC-19 01 24 038).

## References

1. Lu, T., Porter, A. R., Kennedy, A. D., Kobayashi, S. D. & DeLeo, F. R. Phagocytosis and Killing of ***Staphylococcus aureus*** by Human Neutrophils. J Innate Immun 6, 639– 649 (2014).

2. Tamassia, N. et al. Cytokine production by human neutrophils: Revisiting the “dark side of the moon”. Eur J Clin Investigation 48, e12952 (2018).

3. Yang, H. et al. Neutrophil CD16b crosslinking induces lipid raft-mediated activation of SHP-2 and affects cytokine expression and retarded neutrophil apoptosis. Experimental Cell Research 362, 121–131 (2018).

4. Golay, J. et al. Human neutrophils express low levels of FcγRIIIA, which plays a role in PMN activation. Blood 133, 1395–1405 (2019).

5. Koenderman, L. Inside-Out Control of Fc-Receptors. Front. Immunol. 10, 544 (2019).

6. Rivas-Fuentes, S., García-García, E., Nieto-Castañeda, G. & Rosales, C. Fcγ receptors exhibit different phagocytosis potential in human neutrophils. Cellular Immunology 263, 114–121 (2010).

7. Selvaraj, P., Fifadara, N., Nagarajan, S., Cimino, A. & Wang, G. Functional Regulation of Human Neutrophil Fc ⍰ Receptors. 11.

8. Cid, J., Aguinaco, R., Sánchez, R., García-Pardo, G. & Llorente, A. Neutrophil CD64 expression as marker of bacterial infection: A systematic review and meta-analysis. Journal of Infection 60, 313–319 (2010).

9. Wang, X. et al. Neutrophil CD64 expression as a diagnostic marker for sepsis in adult patients: a meta-analysis. Crit Care 19, 245 (2015).

10. Bruhns, P. et al. Specificity and affinity of human Fcγ receptors and their polymorphic variants for human IgG subclasses. Blood 113, 3716–3725 (2009).

11. Patel, K. R., Roberts, J. T. & Barb, A. W. Multiple Variables at the Leukocyte Cell Surface Impact Fc γ Receptor-Dependent Mechanisms. Front. Immunol. 10, 223 (2019).

12. Alemán, O. R., Blanco-Camarillo, C., Naranjo-Pinto, N., Mora, N. & Rosales, C. Fc gamma receptors activate different protein kinase C isoforms in human neutrophils. Journal of Leukocyte Biology qiaf019 (2025) doi:10.1093/jleuko/qiaf019.

13. Fernandes, M. J. G., Lachance, G., Paré, G., Rollet-Labelle, E. & Naccache, P. H. Signaling through CD16b in human neutrophils involves the Tec family of tyrosine kinases. Journal of Leukocyte Biology 78, 524–532 (2005).

14. Fernandes, M. J. G. et al. CD16b associates with high-density, detergent-resistant membranes in human neutrophils. Biochemical Journal 393, 351–359 (2006).

15. Jongstra-Bilen, J., Harrison, R. & Grinstein, S. Fcγ-receptors Induce Mac-1 (CD11b/CD18) Mobilization and Accumulation in the Phagocytic Cup for Optimal Phagocytosis. Journal of Biological Chemistry 278, 45720–45729 (2003).

16. Urbaczek, A. C. et al. Influence of FcγRIIIb polymorphism on its ability to cooperate with FcγRIIa and CR3 in mediating the oxidative burst of human neutrophils. Human Immunology 75, 785–790 (2014).

17. Zhou, M. & Brown, E. J. CR3 (Mac-l, CgM2, CDllb/CD18) and Fc /RIII Cooperate in Generation of a Neutrophil Respiratory Burst: Requirement for FcTRII and Tyrosine Phosphorylation. The Journal of Cell Biology 125, 10 (1994).

18. Fromont, P. et al. Frequency of the polymorphonuclear neutrophil Fc gamma receptor III deficiency in the French population and its involvement in the development of neonatal alloimmune neutropenia. Blood 79, 2131–2134 (1992).

19. Wagner, C. & Hänsch, G. M. Genetic deficiency of CD16, the low affinity receptor for immunoglobulin G, has no impact on the functional capacity of polymorphonuclear neutrophils. Eur J Clin Investigation 34, 149–155 (2004).

20. Riera, N. E. et al. [Neutrophils without CD16b receptors]. Medicina (B Aires) 69, 442– 446 (2009).

21. Minguela, A. et al. Low affinity immunoglobulin gamma Fc region receptor III B (FcγRIIIB, CD16B) deficiency in patients with blood and immune system disorders. Br J Haematol 195, 743–747 (2021).

22. Kremserová, S., Kocurková, A., Chorvátová, M., Klinke, A. & Kubala, L. Myeloperoxidase Deficiency Alters the Process of the Regulated Cell Death of Polymorphonuclear Neutrophils. Front. Immunol. 13, 707085 (2022).

23. Melnik, Z. et al. A rare case of acquired myeloperoxidase deficiency. eJHaem 2, 293– 294 (2021).

24. Salmon, J. E., Brogle, N. L., Edberg, J. C. & Kimberly, R. P. Fc gamma receptor III induces actin polymerization in human neutrophils and primes phagocytosis mediated by Fc gamma receptor II. The Journal of Immunology 146, 997–1004 (1991).

25. Jaumouillé, V. & Waterman, C. M. Physical Constraints and Forces Involved in Phagocytosis. Front. Immunol. 11, 1097 (2020).

26. Li, Z. et al. Aging-Impaired Filamentous Actin Polymerization Signaling Reduces Alveolar Macrophage Phagocytosis of Bacteria. The Journal of Immunology.

27. Negoro, P. E. et al. Spleen Tyrosine Kinase Is a Critical Regulator of Neutrophil Responses to *Candida* Species. mBio 11, e02043–19 (2020).

28. Karlsson, A., Nixon, J. B. & McPhail, L. C. Phorbol myristate acetate induces neutrophil NADPH-oxidase activity by two separate signal transduction pathways: dependent or independent of phosphatidylinositol 3-kinase. Journal of Leukocyte Biology 67, 396–404 (2000).

29. Nanì, S. et al. Src Family Kinases and Syk Are Required for Neutrophil Extracellular Trap Formation in Response to β-Glucan Particles. J Innate Immun 7, 59–73 (2015).

30. Zhu, X. et al. Scavenger Receptor Function of Mouse Fcγ Receptor III Contributes to Progression of Atherosclerosis in Apolipoprotein E Hyperlipidemic Mice. The Journal of Immunology 193, 2483–2495 (2014).

31. Romee, R. et al. NK cell CD16 surface expression and function is regulated by a disintegrin and metalloprotease-17 (ADAM17). Blood 121, 3599–3608 (2013).

32. Wang, Y. et al. ADAM17 cleaves CD16b (FcγRIIIb) in human neutrophils. Biochimica et Biophysica Acta (BBA) - Molecular Cell Research 1833, 680–685 (2013).

33. Huot, S., Laflamme, C., Fortin, P. R., Boilard, E. & Pouliot, M. IgG aggregates rapidly upregulate FcgRI expression at the surface of human neutrophils in a FcgRII dependent fashion: A crucial role for FcgRI in the generation of reactive oxygen species. FASEB j. 34, 15208–15221 (2020).

34. Flesch, B. K., Achtert, G. & Neppert, J. Inhibition of monocyte and polymorphonuclear granulocyte immune phagocytosis by monoclonal antibodies specific for FcgRI, II and III.

35. Andrews, K., Abdelsamed, H., Yi, A.-K., Miller, M. A. & Fitzpatrick, E. A. TLR2 Regulates Neutrophil Recruitment and Cytokine Production with Minor Contributions from TLR9 during Hypersensitivity Pneumonitis. PLoS ONE 8, e73143 (2013).

36. Fang, L., Wu, H.-M., Ding, P.-S. & Liu, R.-Y. TLR2 mediates phagocytosis and autophagy through JNK signaling pathway in Staphylococcus aureus-stimulated RAW264.7 cells. Cellular Signalling 26, 806–814 (2014).

37. Beutler, B., Du, X. & Poltorak, A. Identification of Toll-like receptor 4 (Tlr4) as the sole conduit for LPS signal transduction: genetic and evolutionary studies.

38. Rittirsch, D. et al. Cross-Talk between TLR4 and FcγReceptorIII (CD16) Pathways. PLoS Pathog 5, e1000464 (2009).

39. Shang, L. et al. Selective Antibody Intervention of Toll-like Receptor 4 Activation through Fc ⍰ Receptor Tethering. 289, (2014).

40. Schnoor, M., Vadillo, E. & Guerrero-Fonseca, I. M. The extravasation cascade revisited from a neutrophil perspective. Current Opinion in Physiology 19, 119–128 (2021).

41. De Haas, M., Kleijer, M., Van Zwieten, R., Roos, D. & Von Dem Borne, A. Neutrophil Fc gamma RIIIb deficiency, nature, and clinical consequences: a study of 21 individuals from 14 families. Blood 86, 2403–2413 (1995).

42. Flesch, B. K., Achtert, G., Bauer, F. & Neppert, J. The NA“null” phenotype of a young man is caused by an FcgRIIIB gene deficiency while the products of the neighboring FcgRIIA and FcgRIIIA genes are present.

43. Kiefer, F. et al. The Syk Protein Tyrosine Kinase Is Essential for Fc⍰ Receptor Signaling in Macrophages and Neutrophils. MOL. CELL. BIOL. 18, (1998).

44. Tabata, H., Morita, H., Kaji, H., Tohyama, K. & Tohyama, Y. Syk facilitates phagosome-lysosome fusion by regulating actin-remodeling in complement-mediated phagocytosis. Sci Rep 10, 22086 (2020).

45. De Castro, R. O., Zhang, J., Jamur, M. C., Oliver, C. & Siraganian, R. P. Tyrosines in the Carboxyl Terminus Regulate Syk Kinase Activity and Function. Journal of Biological Chemistry 285, 26674–26684 (2010).

46. Benavente, M. C. R. et al. Distinct CD16a features on human NK cells observed by flow cytometry correlate with increased ADCC. Sci Rep 14, 7938 (2024).

47. Yeap, W. H. et al. CD16 is indispensable for antibody-dependent cellular cytotoxicity by human monocytes. Sci Rep 6, 34310 (2016).

48. Sendler, A. Neutrophils Express the High Affinity Receptor for IgG (FcyRI, CD64) After In Vivo Application of Recombinant Human Granulocyte Colony-Stimulating Factor.

49. Fossati, G., Moots, R. J., Bucknall, R. C. & Edwards, S. W. Differential role of neutrophil Fc? receptor IIIB (CD16) in phagocytosis, bacterial killing, and responses to immune complexes. Arthritis & Rheumatism 46, 1351–1361 (2002).

50. Alemán, O. R., Mora, N., Cortes-Vieyra, R., Uribe-Querol, E. & Rosales, C. Differential Use of Human Neutrophil Fc γ Receptors for Inducing Neutrophil Extracellular Trap Formation. Journal of Immunology Research 2016, 1–17 (2016).

51. Hundt, M. & Schmidt, R. E. The glycosylphosphatidylinositol linked Fcγ receptor III represents the dominant receptor structure for immune complex activation of neutrophils. Eur J Immunol 22, 811–816 (1992).

52. Tsujimoto, H. et al. NEUTROPHIL ELASTASE, MIP-2, AND TLR-4 EXPRESSION DURING HUMAN AND EXPERIMENTAL SEPSIS: Shock 23, 39–44 (2005).

